# Checkpoint-blocker induced autoimmunity is associated with pretreatment T cell expression profiles and favourable outcome in melanoma

**DOI:** 10.1101/2020.06.23.20138594

**Authors:** W. Ye, A Olsson-Brown, R. A. Watson, V. T. F. Cheung, R. D. Morgan, I. Nassiri, R. Cooper, C.A. Taylor, O. Brain, R. N. Matin, N. Coupe, M. R. Middleton, M. Coles, J.J. Sacco, M. J. Payne, B. P. Fairfax

## Abstract

1

**Background:** Immune checkpoint blockers (ICBs) activate CD8^+^ T cells to elicit anti-cancer activity but frequently lead to immune-related adverse events (irAEs). The relationship of irAE with baseline parameters and clinical outcome is unclear. We investigated associations between irAE development, CD8^+^ T cell receptor diversity and expression and clinical outcome in a non-trial setting.

**Methods:** Patients ≥18 years old with metastatic melanoma (MM) receiving combination ICB (ipilimumab plus nivolumab – cICB, n=60) or single-agent ICB (nivolumab/pembrolizumab – sICB, n=78) were prospectively recruited. We retrospectively evaluated the impact of irAEs on survival. This analysis was repeated in an independent cohort of MM patients treated at a separate institution (n=210, cICB:74, sICB:136). We performed RNA sequencing of CD8^+^ T cells isolated from patients prior to treatment, analysing T cell receptor clonality differential transcript expression according to irAE development.

**Results:** 48.6% of patients experienced treatment-related irAEs within the first 5 cycles of treatment. Development of irAE prior to the 5^th^ cycle of ICB was associated with longer progression-free and overall survival (PFS, OS) in the primary cohort (log-rank test, PFS: *P*=0.00034; OS: *P*<0.0001), replicated in the secondary cohort (OS: *P*=0.00064). Across cohorts median survival for those patients not experiencing irAE was 14.4 (95% CI:9.6-19.5) months vs not-reached (95% CI:28.9 - Inf), *P*=3.0×10^−7^. Pre-treatment performance status and neutrophil count, but not BMI, were additional predictors of clinical outcome. Analysis of CD8^+^ T cells from 128 patients demonstrated irAE development was associated with increased T cell receptor diversity post-treatment (*P*=4.3×10^−5^). Development of irAE in sICB recipients was additionally associated with baseline differential expression of 224 transcripts (FDR<0.1), enriched in pro-inflammatory pathway genes including *CYP4F3* and *PTGS2*.

**Conclusions:** Early irAE development post-ICB is strongly associated with favourable survival in MM and increased diversity of peripheral CD8^+^ T cell receptors after treatment. irAE post-sICB is associated with pre-treatment upregulation of inflammatory pathways, indicating irAE development may reflect baseline immune activation states.

**Key message:** Immune-related adverse events (irAEs) commonly occur in patients with metastatic melanoma treated with immune checkpoint blockade (ICB) therapy. In real world setting we find development of early irAEs post-ICB treatment is associated with survival benefit, indicative of a shared mechanism with anti-tumour efficacy. CD8^+^ T cells from patients who develop irAE show increased receptor diversity, and pre-treatment samples from patients who develop irAE post single-agent anti-PD1 show over-expression of inflammatory pathways, indicating baseline immune state can determine irAE development.

## 2 Introduction

The introduction of immune checkpoint blockade (ICB) therapy into clinical practice has transformed the outlook for metastatic melanoma (MM) patients. Current ICB standard of care consists of either monotherapy with anti-PD1 agents (pembrolizumab or nivolumab, sICB), or combined anti-CTLA-4/ anti-PD1 (ipilimumab and nivolumab, cICB). Nivolumab and pembrolizumab block T cell programmed death 1 (PD1) receptor, preventing ligation by the ligands PDL1 and PDL2 which are frequently over-expressed in tumour and stroma [1]. Ipilimumab is an anti-CTLA-4 monoclonal antibody that enhances effector T cell activity by antagonising the homeostatic function of CTLA-4 which is induced upon T-cell antigen presentation [2]. Whereas untreated MM is associated with a 5-year overall survival (OS) of approximately 10% [3], the 5-year OS following anti-PD1 monotherapy in treatment-naive patients is 30 – 40% [1] and combination ICB was associated with a median OS exceeding 5 years in the Checkmate 067 study, with a subset of patients potentially cured [4].

A key concern with ICB is the high incidence of immune-related adverse events (irAEs). Notably, compared to patients receiving sICB, patients receiving cICB show increased incidence and severity of irAEs [5]. IrAEs can be challenging to manage, requiring treatment interruption or discontinuation and systemic immunosuppression. At present, it remains controversial as to whether the development of irAEs is associated with survival. Several retrospective studies have observed an association between the development of irAEs and improved treatment response and/or survival in MM patients treated with ICB, suggesting that reduced tolerance to self-antigens and reduced tolerance to tumour antigens are closely linked [5–11]. This is not consistently observed however [12,13], which may be due to confounders including differences between clinical trial and standard clinical populations, divergences from standard dosing and frequent concurrent exposure to other non-standard of care agents such as vaccines.

Data from the clinical setting regarding the prognostic implications of irAE development remain limited. The treatment pathway within the UK National Health Service is standardised, with patients stratified to cICB or sICB depending on clinical features and patient preferences. Patients whose disease progresses on anti-PD1 sICB have the option of targeted therapy if they have activating *BRAF* mutations, or alternatively second-line ipilimumab; whilst patients with disease progression on cICB have no-further standard of care options available. With this in mind, we have assayed the incidence and severity of irAEs in MM patients treated with either cICB, or anti-PD1 sICB across two prospectively recruited cohorts from tertiary UK centres, to explore the question: does the development of irAEs early in ICB therapy impact clinical outcome? We have subsequently explored CD8^+^ T cell RNA sequencing from a subset of the cohort to investigate the relationship between gene expression and irAE development.

## 3 Methods

### 3.1 Patients

Eligible patients were ≥ 18 years with a confirmed diagnosis of MM, and had received at least one cycle of ICB therapy. 138 Patients were recruited prospectively through the Oxford Radcliffe Biobank, from 23 November 2015 to 15 April 2019. 60 Patients received cICB consisting of ipilimumab 3 mg/kg plus nivolumab 1 mg/kg every 3 weeks for a maximum of 4 treatment cycles, followed by maintenance nivolumab 240 mg fortnightly or 480 mg monthly. 78 Patients receiving sICB therapy included either nivolumab 480 mg monthly, or pembrolizumab 2 mg/kg every 3 weeks. In the replication cohort 74 patients were treated at the Clatterbridge Cancer Centre in Liverpool with cICB and 136 with sICB between 1 January 2016 and 7 January 2019. Patients receiving ICB therapy were treated until unacceptable irAE, progressive disease, death, or patient withdrawal.

### 3.2 Study design

Patients provided written consent to participation within the Oxford Radcliffe Biobank (09/H0606/5+5) and this research was approved via applications: OCHRe 16/A019, 18/A064. For the Clatterbridge dataset, research was approved via the HYST study (12/NW/0525) and local audit approval (17-18/40). Data on patient demographic and clinical characteristics, in addition to the type, severity, date of onset and management of irAEs, were collected retrospectively using electronic medical records. Treatment decisions were determined by the treating clinician. IrAEs were reported according to the National Cancer Institute Common Terminology Criteria for Adverse Events (CTCAE) version 4.03 with pneumonitis being diagnosed via CT. We collected efficacy data consisting of radiological response as defined by the Response Evaluation Criteria in Solid Tumours (RECIST) version 1.1 [15], OS and PFS.

### 3.3 Outcomes

We evaluated irAE characteristics, predictors of irAE development, and the OS and PFS in patients who developed early irAEs compared to those who did not. Early irAEs were defined as those from treatment initiation to completion of the 4^th^ cycle of treatment, i.e. occurring prior to cycle 5. We chose this time point as cICB therapy consists of a maximum of 4 cycles of ipilimumab plus nivolumab, prior to commencement of maintenance nivolumab monotherapy. OS was defined as the time from first ICB dose to death from any cause. PFS was defined as the time from first ICB dose to disease progression, as determined by serial cross-sectional imaging, or death.

### 3.4 Statistical analysis

Baseline characteristics were analysed using descriptive statistics. Categorical variables were summarised using frequencies and percentages, and continuous variables using medians and ranges. Given the time-dependent nature of developing irAEs and the susceptibility to guarantee-time bias [15], we also performed a 12 week landmark analysis. Only patients who are alive or have not progressed at 12 weeks are included in the OS and PFS landmark analysis respectively, and patients were grouped according to whether they experienced irAEs prior to 12 weeks. OS and PFS was estimated using Kaplan-Meier analysis, and the log-rank test was used to determine the statistical significance between the curves. Logistic regression was used to determine predictors of developing irAEs. Univariable and multivariable Cox proportional hazards models were used to investigate the association between prognostic factors and survival. *P* < 0.05 was considered statistically significant. All analyses were performed using the survminer [17] and survival [18] packages in R, version 3.5.1.

### 3.5 Sample collection

Patients provided written informed consent to donate samples for analysis to the Oxford Radcliffe Biobank (Oxford Centre for Histopathology Research ethical approval nos. 16/A019 and 18/A064); 30–50□ml blood was collected into EDTA tubes (BD vacutainer system) taken immediately pre-treatment. Peripheral blood mononuclear cells were obtained by density centrifugation (Ficoll Paque). CD8^+^ cell isolation was carried out by positive selection (Miltenyi) according to the manufacturer’s instructions, with all steps performed either at 4°C or on ice. Post-selection cells were spun down and resuspended in 350μl of RLTplus buffer with 1% beta-mercaptoethanol or DTT, and transferred to 2-ml tubes. Samples were stored at −80°C for batched RNA extraction. Homogenization of the sample was carried out using the QIAshredder (Qiagen). The AllPrep DNA/RNA/miRNA kit (Qiagen) was used for RNA extraction. DNase□I was used during the extraction protocol to minimize DNA contamination. RNA was eluted into 35□μl of RNase-free water. The amount of RNA present was quantified by Qubit analysis, and RNA samples stored at − 80°C until ready for sequencing.

### 3.6 Expression analysis

Poly(A) RNA was sequenced on Illumina HiSeq-4000 (75bp paired-end reads) and Illumina Novaseq machines (150bp paired-end reads) both at the Oxford Genome Centre, Wellcome Centre for Human Genetics. Reads were aligned to GCRh38/hg38 using HISAT2, and High-mapping quality reads were selected based on MAPQ score using bamtools. Marking and removal of duplicate reads were performed using picard (v.1.105), and samtools was used to pass through the mapped reads and calculate statistics. 128 high-quality transcriptomes were used for expression analysis of pre-treatment samples. Read count information was generated using HTSeq and DESeq2 [19][20]was used for differential expression analysis, comparing individuals who developed irAE within the initial 5 cycles of treatment versus unaffected. We controlled for sequencer, age and sex in the analysis. Only transcripts with mean of >10 reads were analyzed, using the binomial Wald test with 750□iterations after correcting for size factors and dispersion.

### 3.7 T cell receptor diversity

MiXCR[21] was used to map reads on reference sequences of V, D and J genes, and to quantitate TCR clonotypes from mapped reads using complementarity-determining region 3 (CDR3) gene regions for RNA samples from pre and post first cycle ICB (n=106 paired samples). The non-default partial alignments option (OallowPartialAlignments = true) was applied to preserve partial alignments for the assembly step. Three iterations of read assembly were performed using the assemblePartial setting. Shannon diversity was calculated within the Vegan R package[22].

## 4 Results

### 4.1 Patient characteristics

138 MM patients treated with ICB therapy were prospectively recruited (Table 1). Of 78 patients receiving sICB therapy, 69 received pembrolizumab and 9 received nivolumab. 6 patients had a history of prior autoimmune disease (1 type 1 diabetes, 2 inflammatory bowel disease, 2 thyroid abnormalities, 1 Sjogren’s syndrome). The median number of cycles received per patient was 4 for cICB therapy, and 8 for sICB therapy. The median follow up duration was 12.1 (0.3 – 42.7) months.

**Table 1.** Baseline demographic and clinical characteristics of patients

| Characteristic | Single ICB therapy<br>(n=78) | Combination ICB<br>therapy (n=60) | Complete cohort<br>(n=138) |
| --- | --- | --- | --- |
| Median age (range), years | 74 (29 – 95) | 59 (21 – 74) | 68 (21 – 95) |
| Males, n (%) | 42 (53.8) | 34 (56.7) | 76 (55.1) |
| Median BMI (range) | 28 (21 – 59) | 27 (19 – 43) | 28 (19 – 59) |
| Prior autoimmune disease, n (%) | 4 (5.1) | 2 (3.3) | 6 (4.3) |
| ECOG performance status, n (%) |  |  |  |
| 0 | 29 (37.2) | 47 (78.3) | 76 (55.1) |
| 1 | 35 (44.9) | 10 (16.7) | 45 (32.6) |
| 2 | 12 (15.4) | 3 (5.0) | 15 (10.9) |
| 3 | 1 (1.3) | 0 (0.0) | 1 (0.7) |
| Not reported | 1 (1.3) | 0 (0.0) | 1 (0.7) |
| BRAF status, n (%) |  |  |  |
| Mutation | 19 (24.3) | 23 (38.3) | 42 (30.4) |
| Wild type | 51 (65.4) | 29 (48.3) | 80 (58.0) |
| Not reported | 8 (10.2) | 8 (13.3) | 16 (9.0) |
| Elevated LDH, n (%) | 22 (28.2) | 19 (31.7) | 41 (29.7) |
| Prior systemic treatment, n (%) | 27 (34.6) | 8 (13.3) | 35 (25.4) |
| BRAF inhibitor | 10 (12.8) | 5 (8.3) | 15 (8.7) |
| Ipilimumab | 17 (21.8) | 0 (0.0) | 17 (12.3) |
| Other | 0 (0.0) | 3 (5.0) | 3 (2.2) |

### 4.2 Immune related adverse events

Any grade treatment-related irAEs were reported in 89 (64.5%) patients, of which 74 (53.6%) experienced an irAE prior to cycle 5. Grade 3 or 4 irAEs were reported in 45 (32.6%) patients (Table 2). There were no drug-related deaths. Cutaneous irAEs, colitis and hepatitis occurred early post ICB initiation (median 37, 34 and 49 days respectively). In contrast, gastritis and pneumonitis are late complications, with median time to onset of 378 and 386 days (Supplementary Figure 1a, Table 2).

**Table 2.**
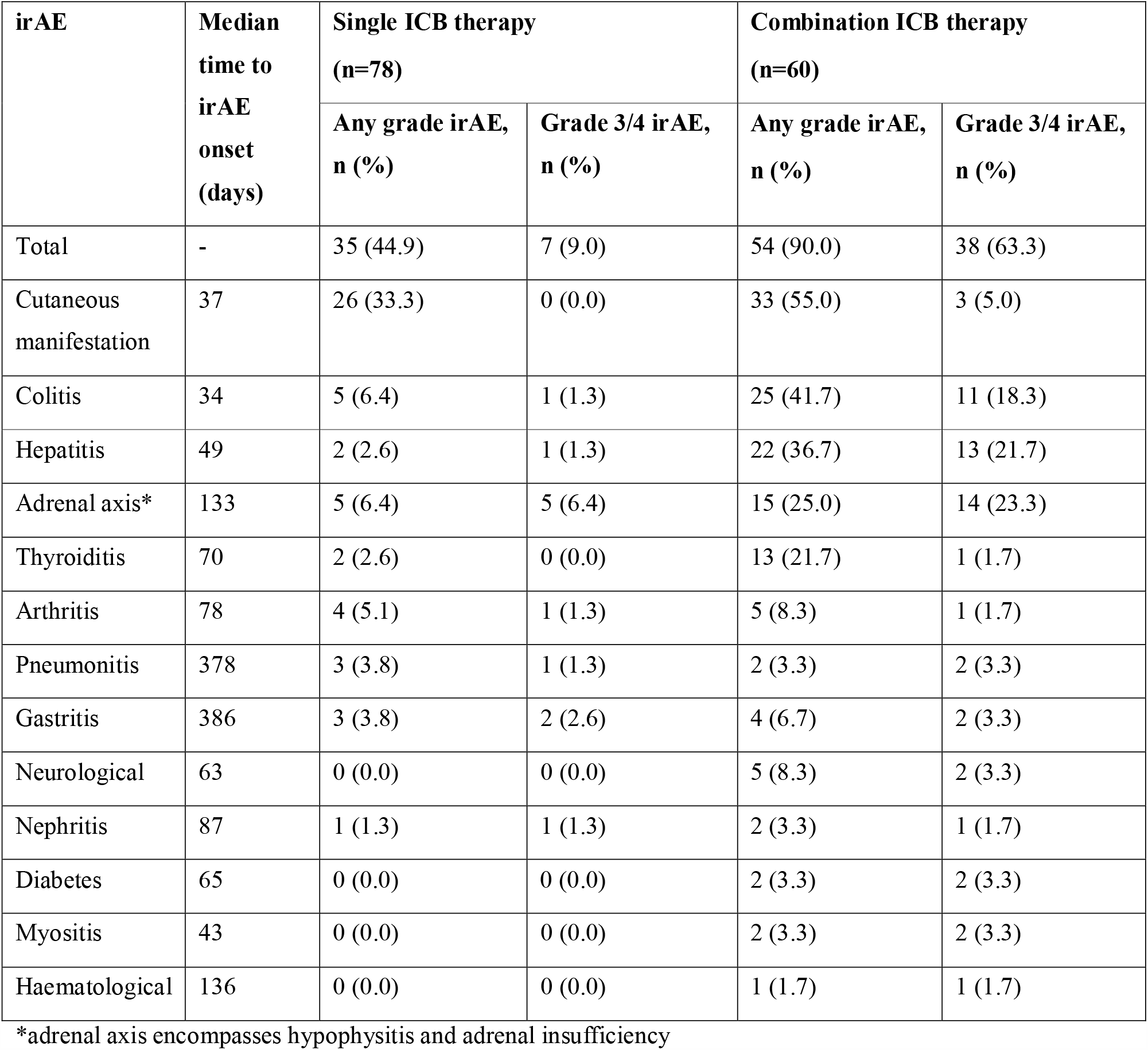
Immune related adverse events (irAEs) across the Oxford cohort

Compared to patients treated with sICB, patients receiving cICB had over two-fold increase in the frequency of any grade irAEs, and more than a four-fold increase in the frequency of Grade 3 or 4 irAEs (Table 2). Multi-organ system irAEs were more common with cICB therapy, with 24 (40%) patients experiencing irAEs affecting 3 or more organs, compared to 3 (4%) patients treated with sICB (Supplementary Figure 1b). Systemic steroids were required in 35/40 (88%) cICB and 10/29 (34%) sICB patients who experienced irAEs. 11 patients had steroid-refractory disease, requiring mycophenolate (4 hepatitis, 1 bullous pemphigoid), infliximab (3 colitis, 1 gastritis), methotrexate (1 arthritis) and hydroxychloroquine (1 arthritis). ICB discontinuation occurred in 30/69 (43%) patients who experienced irAEs (23/40 cICB, 7/29 sICB). Events included colitis (*N* = 7), hepatitis (*N* = 4), cutaneous manifestations (*N* = 3) and hypophysitis (*N* = 3). Neurological irAEs, pneumonitis, autoimmune haemolytic anaemia, diabetes, nephritis and myositis led to ICB discontinuation in 1-2 patients each. In a further 14 patients, ICB therapy was interrupted due to irAEs (11 cICB, 3 sICB). Prior to treatment interruption, the median number of cycles received was 2 and 10 for patients treated with cICB and sICB, respectively.

We used logistic regression to test putative risk factors for irAE development prior to cycle 5, evaluating age, sex, baseline BMI, medical history of autoimmune disease, and type of treatment. We found the only significant clinical predictor of increased likelihood of developing early irAEs being treatment with cICB (OR=19.1, 95% CI 7.1-59.1, *P* =3.8×10^−8^, Supplementary Figure 1c).

### 4.3 Oncological outcomes

Among 138 patients, 58 (42%) experienced a complete or partial response to ICB therapy at the first radiological assessment (3 month CT), whereas 26 patients (18.8%) had stable disease, and 43 (31.2%) had progressive disease. For the remaining 11 (8%) patients, no sequential cross-sectional imaging was available, however 9 (6.5%) patients had clear clinical progression.

The median OS across the cohort was 28.9 months (95% CI 18.7 months – Inf) and the median PFS was 9.0 months (95% CI 5.8-20.5 months). At 1 year, the OS and PFS rates were 72% (95% CI 65 – 81) and 48% (95% CI 40 – 58), respectively. At 2 years, the OS and PFS rates were 56% (95% CI 47 – 67) and 37% (95% CI 28.9 – 48.2), respectively.

Comparing patients who develop an early irAE prior to the 5^th^ cycle of treatment to those who do not, early irAE development was associated with significantly longer OS and PFS (Figure 1a,b, OS log-rank *P* < 0.0001, PFS log-rank *P* = 0.00034). This observation remained significant when stratifying patients according to treatment received (Figure 1c,d, Supplementary Figure 2a,b). Similarly this remained significant when analysis was confined to patients suffering Grade 3 or 4 irAE only (Supplementary Figure 2c,d). To adjust for guarantee-time bias, we performed a landmark analysis including only patients who are alive (*N* = 127) or who had not progressed (*N* = 98) at 12 weeks. We found that the development of an irAE prior to week 12 was associated with significantly improved OS (Supplementary Figure 2e, log-rank *P* =0.0013), however this did not reach significance for PFS (Supplementary Figure 2f, log-rank *P* = 0.099)

**Figure 1.**
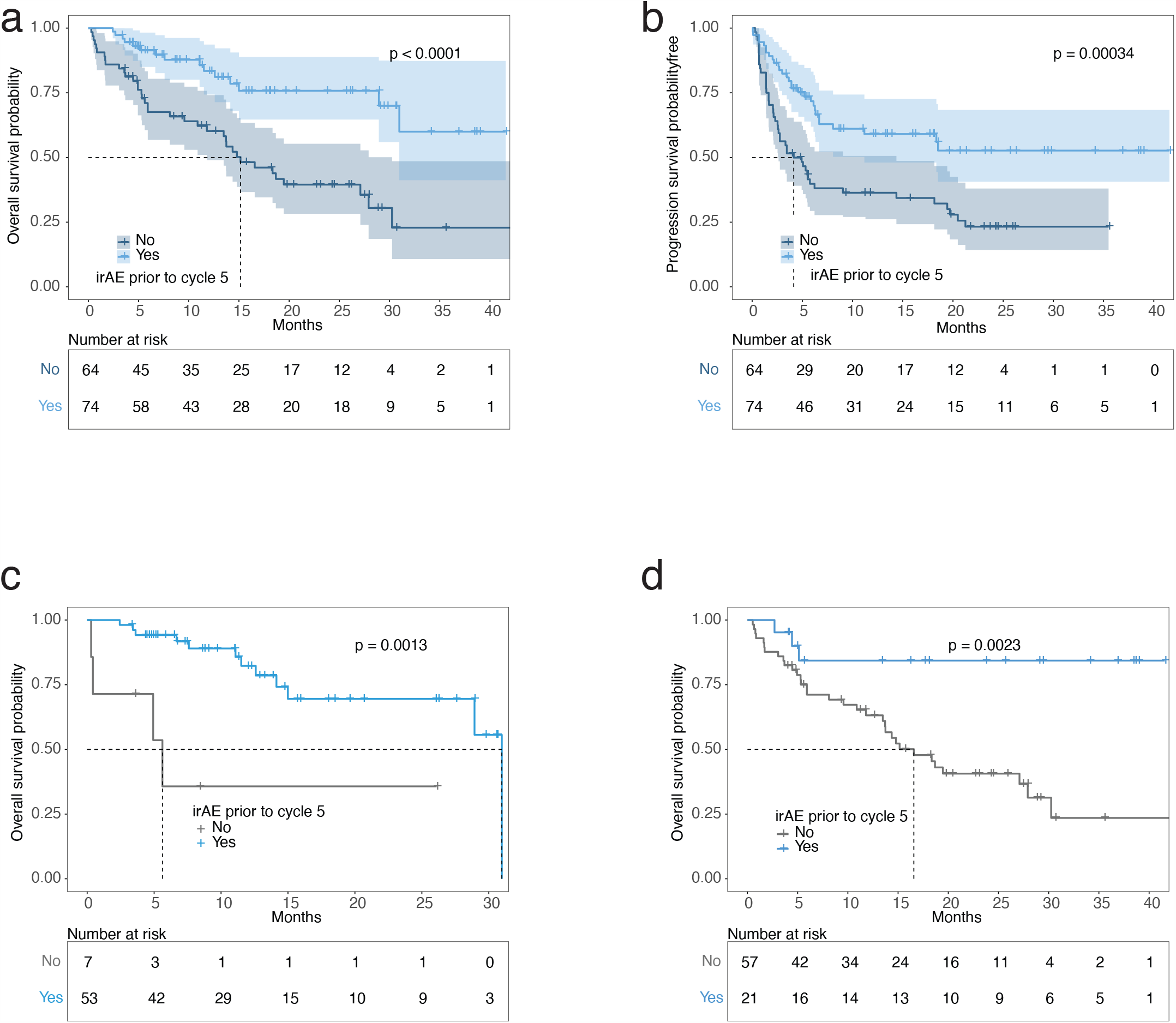
Kaplan Meier curves of overall survival (OS) and progression free survival (PFS) according to development of any grade of irAEs prior to cycle 5. a) OS whole cohort, shaded areas showing 95% CI (n=138), b) PFS whole cohort, shaded areas showing 95% CI, c) Kaplan Meier of OS specific to recipients of cICB therapy (n=60), d) Kaplan Meier of OS specific to recipients of sICB therapy (n=78). All P-values refer to log-rank test.

### 4.4 Independent replication

To explore whether our findings were reproducible in a different institution we repeated the analysis in a separately collected cohort of MM patients receiving ICB based at The Clatterbridge Cancer Centre, Liverpool, consisting of 136 recipients of sICB and 74 recipients of cICB. The cohort had a similar demographic make-up to the Oxford cohort, although the patients who received cICB tended to be older (median age 65 vs. 59, Table 3), whereas the recipients of sICB were younger (median age 68 vs. 74, Table 3). Across this cohort we replicated the observation that irAE during the first 4 cycles of immunotherapy was significantly associated with prolonged overall survival (median 13 (95% CI:8-23) months vs. not reached, *P* =0.00064, Figure 2a). When we assessed each treatment type independently we observed a suggestive but non-significant benefit of irAE within the sICB cohort (median 15 (9-23) vs. 26 (16-Inf) months, P=0.1). Conversely, this observation remained robust within the cICB cohort (median 5 (3-Inf) months vs not reached (20-Inf), *P*=0.00023). When we combined the data from both cohorts early irAE were found to be highly significantly associated with overall survival time (median 14.4 (95% CI:9.6-19.5) months vs not-reached (95% CI:28.9 - Inf), *P*=3.0×10^−7^), Figure 2b), and this remained the case for sICB (median 15.2 (12-23) months vs not-reached (18-Inf), *P*=0.0028) or cICB treatments (median 5 (3-Inf) months vs 31 (28.9 – Inf) months, *P*=3.7×10^−7^).

**Table 3.** Comparison of Oxford and Liverpool cohorts

| Characteristic | sICB<br>(n=136) | P vs.<br>Oxford | cICB (n=74) | P vs.<br>Oxford | All treatments |  |  |
| --- | --- | --- | --- | --- | --- | --- | --- |
|  |  |  |  |  | Liverpool<br>cohort<br>(n=210) | Oxford<br>cohort<br>(n=138) | P |
| Median age<br>(range), years | 68 (28 –<br>89) | P=0.02 | 65 (29 – 87) | P=0.002 | 66 (28 –<br>89) | 68 (21-95) | 0.51 |
| Males, n (%) | 77 (56.6) | P=0.80 | 42 (56.7) | P=1 | 119 (56.7) | 76 (55.1) | 0.77 |
| Median BMI<br>(range) | 28 (21 –<br>59) | P=0.31 | 27 (19 – 43) | P=0.21 | 27 (17-49) | 28 (19 –<br>59) | 0.12 |

**Figure 2.**
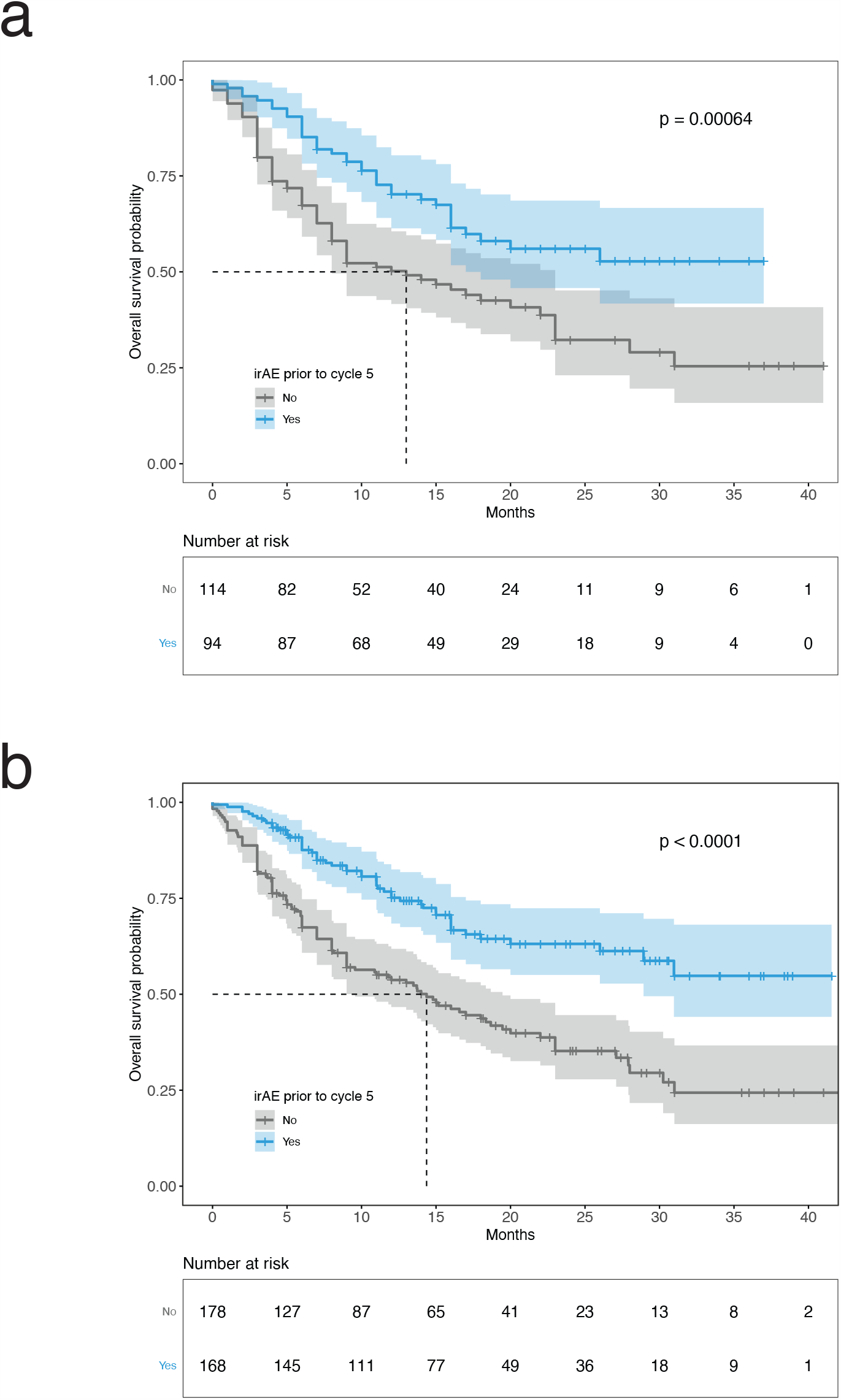
a) Kaplan Meier curves of overall survival (OS) for Liverpool Replication dataset stratified according to development of irAE prior to the fifth cycle of treatment, shaded areas showing 95% CI, (n=218), b) Kaplan Meier curves for combined Oxford and Liverpool datasets (n=346). All P-values refer to log-rank test.

### 4.5 Other variables associated with outcome

We used univariable Cox proportional hazard models to explore the effect of different parameters on OS (Table 4). Development of an irAE prior to cycle 5 and baseline albumin levels were positively associated with significantly improved OS. In contrast, non-cutaneous melanoma subtype, raised performance status, neutrophil count, monocyte count and lactate dehydrogenase levels at baseline were negatively associated with OS (Table 4). Using retrospective trial data from immunotherapy and targeted agents it has recently been shown that a raised BMI in females is associated with a superior clinical outcome[23]. We explored the association between BMI and outcome in both the Oxford and Liverpool cohorts but notably did not see an effect in either univariable or multivariable analyses with clinical outcome, with a trend in both datasets towards raised BMI at treatment start being negatively associated with outcome.

**Table 4.**
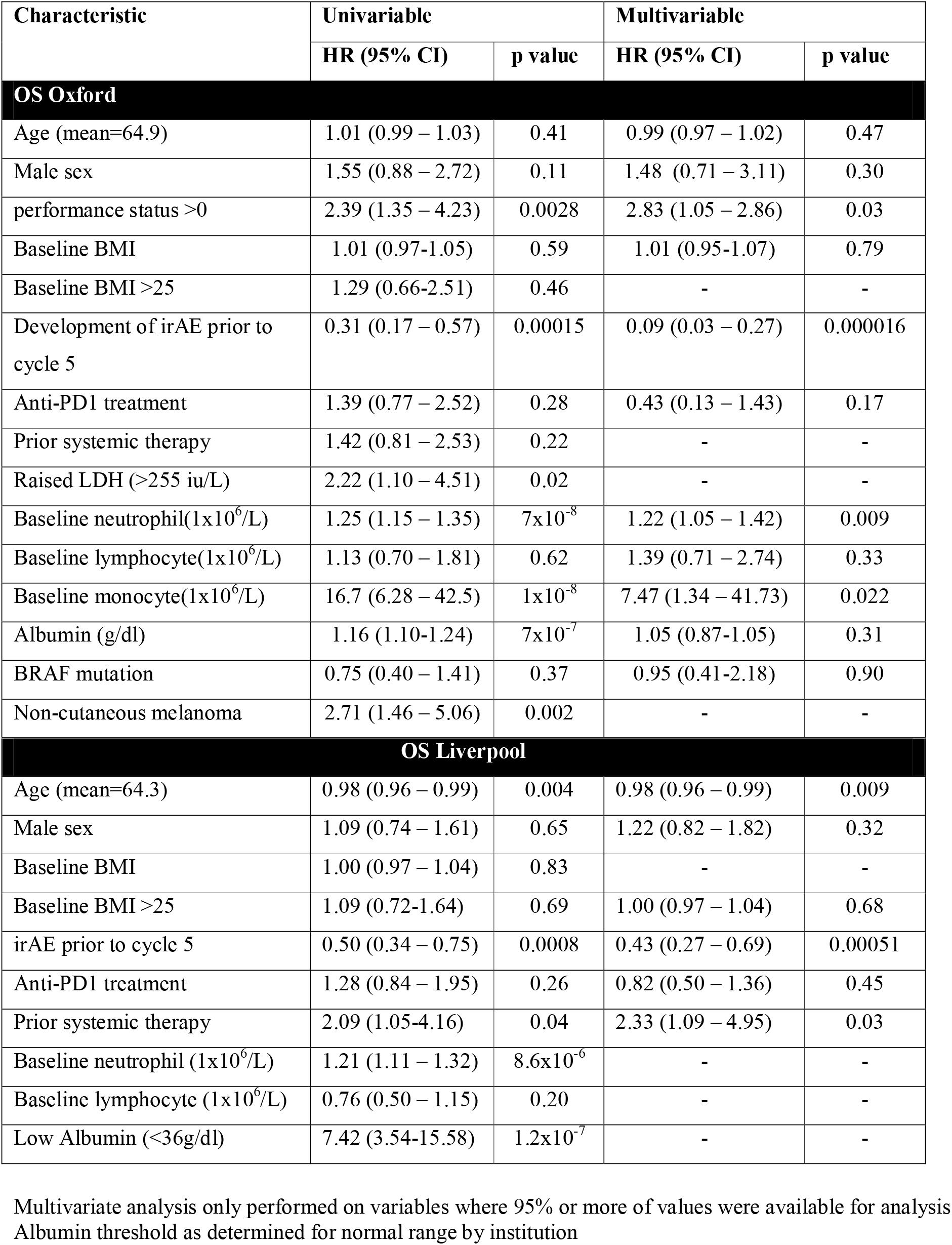
Predictors of oncological outcomes using univariable and multivariable Cox proportional hazards models

**Table 5.** Univariable and multivariable analyses of factors associated with OS - combined datasets

| Characteristic | Univariable |  | Multivariable |  |
| --- | --- | --- | --- | --- |
|  | HR (95% CI) | p value | HR (95% CI) | p value |
| **OS - baseline indices (n=348)** |  |  |  |  |
| Age | 0.99 (0.98 – 1.00) | 0.11 | 0.98 (0.97 – 1.00) | 0.02 |
| Male sex | 1.23 (0.90 – 1.69) | 0.2 | 1.29 (0.93 – 1.79) | 0.13 |
| Baseline BMI | 1.01 (0.98-1.03) | 0.67 | - | - |
| Baseline BMI >25 | 1.13 (0.79-1.60) | 0.51 | 1.10 (0.77 – 1.57) | 0.60 |
| irAE prior to cycle 5 | 0.43 (0.31 – 0.60) | $6.5 \times 10^{-7}$ | 0.38 (0.25 – 0.57) | $2.5 \times 10^{-6}$ |
| Anti-PD1 treatment | 1.33 (0.94 – 1.89) | 0.10 | 0.86 (0.56 – 1.32) | 0.49 |
| Prior systemic therapy | 1.10 (0.68 – 1.79) | 0.69 | 1.14 (0.64 – 2.05) | 0.65 |
| Region (vs. Oxford) | 1.32 (0.95-1.85) | 0.10 | 1.31 (0.88-1.95) | 0.19 |
| **OS - including biochemical and haematological indices (n=281)** |  |  |  |  |
| Age | - | - | 0.99 (0.97 – 1.00) | 0.14 |
| Male sex | - | - | 1.35 (0.91 – 1.99) | 0.14 |
| Baseline BMI >25 | - | - | 0.96 (0.61 – 1.50) | 0.85 |
| irAE prior to cycle 5 | - | - | 0.36 (0.23 – 0.58) | $1.6 \times 10^{-5}$ |
| Anti-PD1 treatment | - | - | 1.25 (0.74 – 2.10) | 0.41 |
| Prior systemic therapy | - | - | 1.22 (0.66 – 2.25) | 0.52 |
| Baseline neutrophil | 1.22 (1.15 – 1.29) | $1.2 \times 10^{-11}$ | 1.17 (1.09-1.27) | $3.4 \times 10^{-5}$ |
| Baseline lymphocyte | 1.11 (0.66 – 1.23) | 0.53 | 0.96 (0.69-1.33) | 0.80 |
| Low Albumin (per centre) | 4.7 (2.87-7.70) | $8.1 \times 10^{-10}$ | 3.68 (1.96-6.91) | $5.3 \times 10^{-5}$ |
| Region (vs. Oxford) | - | - | 1.55 (0.41-1.01) | 0.06 |

Multivariate analysis only performed on variables where 95% or more of values were available for analysis Albumin threshold as determined for normal range by institution

Using multivariable models to explore the interaction between these different factors, we found that development of an irAE prior to cycle 5, monocyte count, performance status and neutrophil count were nominally associated with OS (Table 4), although irAE remained the only significant predictor after correcting for multiple testing (Figure 3a, HR 0.09, 95% CI: 0.03 – 0.27, (Benjamini Hochberg corrected) *FDR*=0.0002). We had access to more limited clinical data from the Liverpool cohort, but in the subset of individuals with neutrophil counts available we replicated the negative association with baseline neutrophil count and clinical outcome (Table 4).

**Figure 3.**
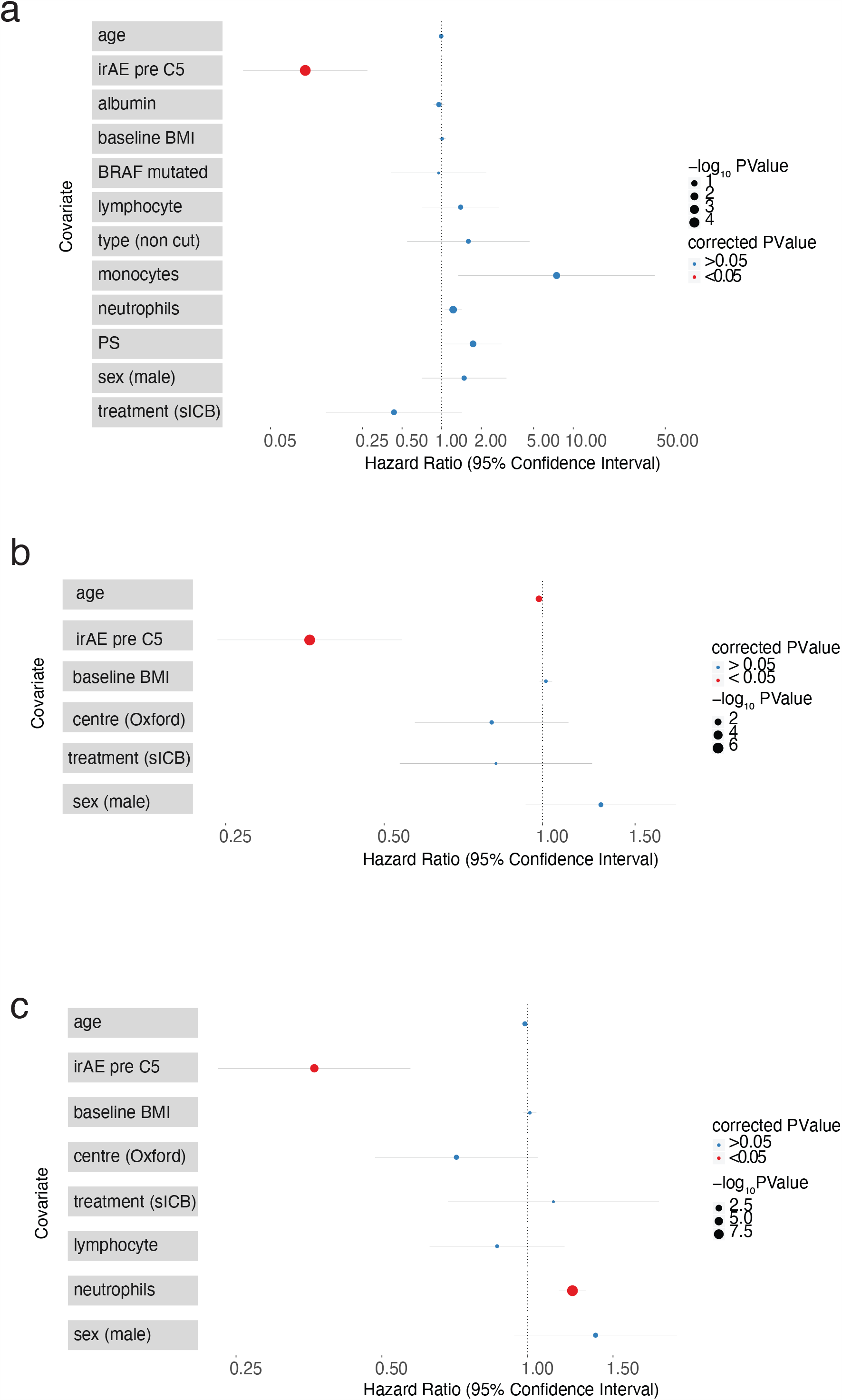
a) Results from multivariable Cox Proportional Hazard analysis of factors associated with OS in Oxford cohort (n=138), P values corrected for multiple testing cohort, b) Factors associated with OS across combined Oxford and Liverpool datasets where data available for all individuals (n=348), c) Factors associated with OS across combined Oxford and Liverpool datasets where data available including cell counts (n=272).

Performing multivariable Cox proportional hazard models on the combined datasets from both institutions (n=348) we again observed a protective effect of irAE (HR 0.35, 95% CI:0.24-0.53, *P*=6.8 x10^−7^) and a weaker effect of age (reduction in HR 0.98, 95% CI:0.97-0.99, *P*=0.02, Figure 3b). Across both sites we had data including baseline lymphocyte and neutrophil counts for 272 individuals. Analysis of these combined data demonstrated that baseline neutrophil count (HR 1.24 per unit increase, 95% CI:1.16-1.32, *FDR*=1.3×10^−9^) and development of irAE prior to cycle 5 (HR 0.36, 95% CI:0.23-0.57, *FDR*=5.4×10^−5^) were associated with clinical outcome (Figure 3c).

### 4.6 Association of irAE development with CD8^+^ T Cell Receptor diversity and baseline gene expression

Identification of markers predictive of irAE is of high interest to immuno-oncology. There is growing evidence that variation in propensity to irAE might be ascertainable by analysis of blood samples[24]. We have previously shown that analysis of peripheral CD8^+^ T cell transcriptomics can provide insights into treatment outcome[25]. Given the association between clinical outcome and irAE development we sought to further investigate the relationship between irAE development and features of peripheral CD8^+^ T cells. We analysed T Cell Receptor (TCR) diversity from CD8^+^ T cells from pre (day 0) and post-treatment (day 21) samples from 128 of the patients in the Oxford cohort. Analysis of the Shannon Diversity index, a marker of abundance and evenness, of TCR demonstrated that patients who proceeded to develop irAE within the first five cycles of treatment had significantly greater CD8^+^ T cell TCR diversity on day 21 post treatment (Figure 4a). Notably we found a strong inverse-relationship between age and diversity, and this was reflected in a difference between the pre-treatment diversity in recipients of sICB and cICB where recipients in the Oxford cohort differed in age. To further explore the relationship between irAE development and CD8^+^ T cell TCR diversity we fitted a linear model, taking into account treatment type, patient age, day 0 and day 21 CD8^+^ TCR diversity, sex and baseline monocyte counts. This demonstrated that, in addition to treatment type, TCR diversity on day 21 was a key predictor of irAE (*P*=0.038, Figure 4b).

**Figure 4.**
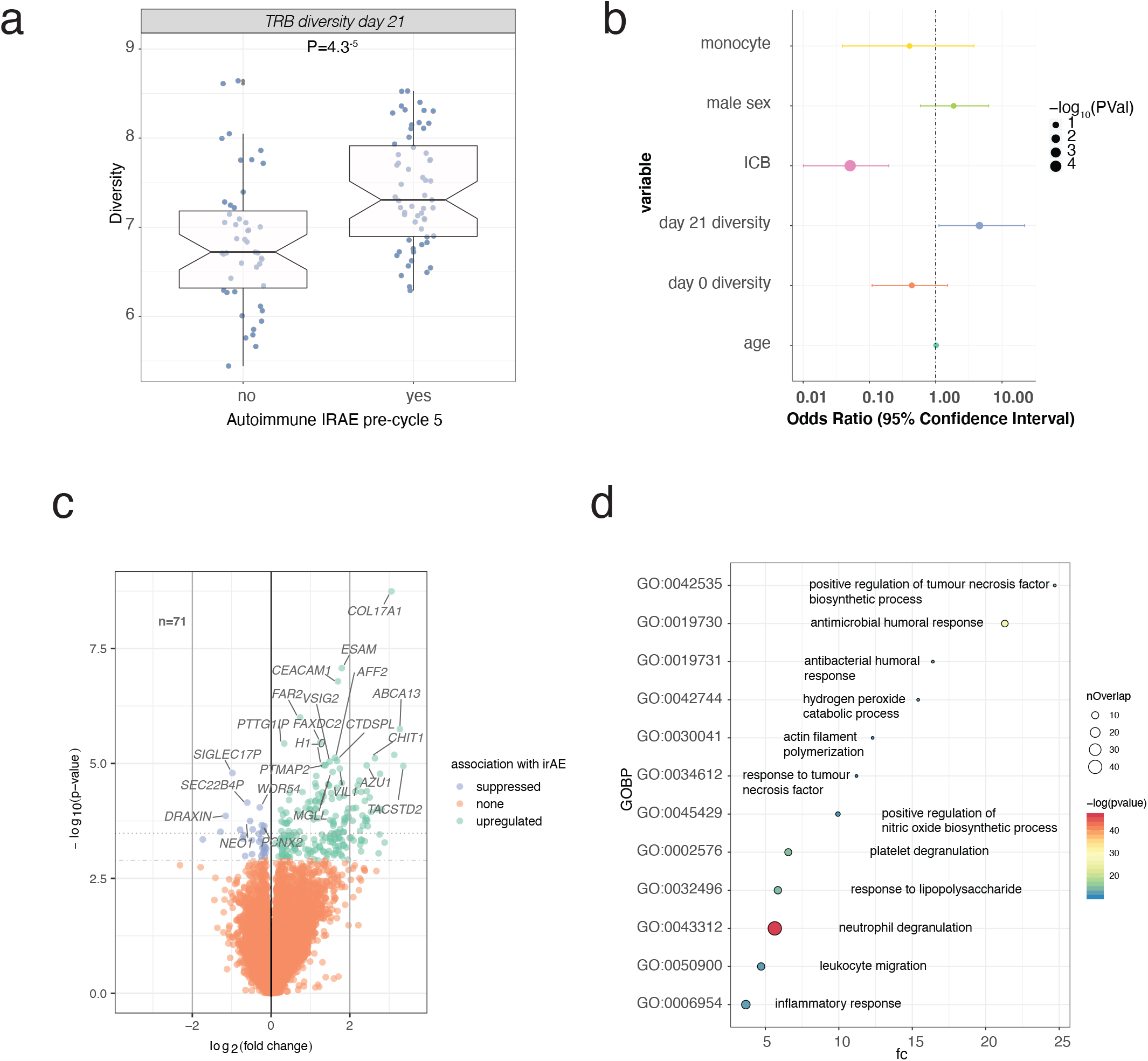
a) The diversity of T Cell receptor beta chain, calculated from CD8^+^ TCR data mapped from RNA on day 21 of ICB treatment was significant greater in those who developed irAE (student T test, n=106), b) Results from multivariable logistic regression analysis demonstrating significant association with both ICB type (reference cICB) and day 21 TCR diversity with the development of irAE, c) Volcano plot of differentially expressed transcripts from pre-treatment CD8+ T cells from sICB recipients comparing those who did and did not proceed to develop irAE within the first 5 cycles of treatment (n=71, 116 transcripts at FDR=0.05 and 224 transcripts at FDR=0.1) d) Go Ontology Biological Process (GOBP) pathway analysis of genes differentially associated with development from 4c. Here the most significant 12 pathways are shown (y axis: GOBP code, x axis: fold change enrichment).

### 4.7 Association with CD8^+^ T cell baseline gene expression

To explore the relevance of baseline transcriptomics to irAE development we analysed gene expression in CD8^+^ T cells isolated from ICB recipients immediately prior to treatment. Whilst we saw limited significant variation in pre-treatment samples according to development of irAE post cICB (data not shown), analysis of CD8^+^ T cell expression data of sICB recipients (n=71), of whom 24% (17/71) developed irAE prior to the fifth cycle of treatment, identified 224 transcripts (*FDR*<0.1, Supplementary Data 1) differentially expressed according to subsequent development of an irAE (Figure 4c). Notable upregulated genes included several involved in arachidonic acid metabolism and eicosanoid synthesis including *CYP4F3*, encoding Leukotriene-B(4) omega-hydroxylase 2 (FDR 0.014), and *PTGS2*, encoding prostaglandin-endoperoxide synthase 2 (FDR 0.021). Pathway analysis of transcripts upregulated according to subsequent irAE development showed 58 pathways where there was enrichment of genes upregulated in patients who developed irAE (*FDR*<0.05, Supplementary Data 2). Notably the greatest fold enrichment was observed in genes involved in TNF synthesis (GO:0042535, 24.7 fold-change, *P*=2.5×10^−7^), a key cytokine putatively involved in irAE development (Figure 4d, Supplementary Data 2). These data demonstrate a baseline tendency to inflammation before treatment in individuals who develop irAE post sICB, suggesting that the pre-treatment immune state sensitizes to irAE development.

## 5 Discussion

Whilst ICB therapy constitutes the main treatment for MM, it frequently results in irAE development. Consistent with the literature [5], we find a markedly higher incidence of irAEs in patients treated with cICB compared to sICB therapy. Moreover, irAEs secondary to cICB are typically more severe, with a greater proportion of patients experiencing multi-organ involvement. Neither sex nor prior autoimmune history influenced the development of early irAEs in our cohort, whereas in the general population, autoimmune diseases have a higher prevalence in females and in those with a history of autoimmunity [26]. Although we were underpowered to exclude absence of effect of prior autoimmune history, the lack of sex effect suggests classical risk factors for autoimmune disease may be of less relevance to the development of irAEs. Further studies are important to ascertain whether this observation remains robust. We could not replicate the reported association between pre-treatment BMI and clinical outcome [23] in either the Oxford or Liverpool datasets in either sex and there was no association between BMI and tendency to irAEs. Our data is notable in that the BMI was collected prospectively, and it did not incorporate targeted agents or immunotherapy and chemotherapy regimens together. Whilst further prospective data is required, these results suggest the putative link between BMI and favourable outcome in ICB is not clear-cut and prospective studies analysing this would be helpful.

Awareness of the different irAEs and their clinical characteristics is paramount to their successful management in the clinical setting. As previously described, we observe that distinct irAEs have a different median time-to-onset post ICB therapy initiation, which may reflect diverging mechanisms of development. Cutaneous irAEs, colitis and hepatitis typically occur early, whilst late complications include gastritis and pneumonitis. The timescales within our cohort broadly corroborate those reported in previous studies [9,19], except pneumonitis which occurs at a median time of 378 days within our cohort compared to 62 days in a previous study [10]. Despite the diversity of irAEs that occur with ICB therapy, most can be managed symptomatically or with corticosteroids, and only a small number require other systemic therapies.

Whilst association between development of irAEs and oncological response to ICBs has been described previously, it has been predominantly in the trial setting [6,7]. Previous studies have demonstrated vitiligo development to be linked to an objective response to ICB therapy [11,12], although the association between vitiligo and good clinical outcome in melanoma is known outside ICB treatment. A link between irAE and survival benefit in ICB therapy has also been reported in non-small cell lung cancer and therefore may not be specific to melanoma [28]. However, not all available evidence support the link between irAE and improved survival with retrospective studies of single agent Ipilimumab response and nivolumab responses failing to observe an association [9,12,13].

With data from a standard-of-care non-trial dataset, we clearly demonstrate that development of irAE of any grade is associated with substantially improved OS, across univariable and multivariable Cox models. This observation remains significant in the 12-week landmark analysis, ruling out a guarantee-time bias where patients died before being able to develop irAE. In contrast, the association between early irAE and improved PFS was significant across univariable and multivariable analyses, but not in the 12-week landmark analysis. Importantly, we were able to replicate these observations in an independent cohort, although here, upon analysis of individual treatments the effect only remained significant in recipients of cICB, although a strong trend to this effect in the sICB group existed with more marked variation in response. Notably, there were significant differences between the Oxford and Liverpool cohorts, with Liverpool sICB recipients tending to be younger and Liverpool cICB recipients significantly older on average than those in Oxford. When we combined the datasets in multivariable analyses however, development of irAE was highly associated with outcome, even when taking treatment type into account. Similarly, neutrophil count at start of treatment was strongly negatively associated with outcomes, indicating baseline myeloid proliferation leading to neutrophilia is a clear poor prognostic marker of response.

The incorporation of the TCR diversity data and transcriptomic analysis from a reasonably powered dataset (n=128 individuals) demonstrated that higher diversity of TCR post treatment was found amongst those who developed irAE. Notably, TCR diversity is associated with age, as was the type of treatment patients were placed on in Oxford. However, when accounting for treatment type, age, sex, monocyte count, and baseline diversity we still found that development of irAE remained associated with day 21 TCR diversity. A greater TCR diversity equates to increased ability to recognise diverse antigens, and thus an association with propensity to irAEs may be anticipated.

Whereas we did not note a strong association between baseline CD8^+^ cell gene expression and irAE development in recipients of cICB, across sICB recipients who developed irAEs, increased expression of inflammatory pathway mediators was noted in the pre-treated state. cICB leads to markedly greater changes in gene expression in CD8^+^ T cells than sICB[25] and we postulate that with such broad activation, baseline variation in expression is of less importance compared to the larger effect of treatment. Within the sICB recipients who develop irAE we suspect that the enrichment in neutrophil and platelet activation pathways in CD8^+^ T cells at baseline may reflect increased intercellular adherence in these samples with platelet adherence to immunoprecipitated T cells, a common finding in activated T cells from chronic viral infection[29], which is postulated to aid CD8^+^ recruitment to sites of injury. Increased cellular adherence indicates predisposal to activation, and this is supported by enrichment of numerous inflammatory mediator pathways including hydrogen peroxide catabolic process (GO:0042744), positive regulation of tumour necrosis factor biosynthetic process (GO:0042535) and positive regulation of nitric oxide biosynthetic process (GO:0045429). Thus our data suggests the propensity to develop irAE to sICB is in part due to the baseline CD8^+^ T cell activation, which can be predicted and may reflect pre-treatment anti-tumour responses.

It can be inferred that because irAE were managed as per standard, frequently with steroids or other immunosuppressants, these treatments are unlikely to adversely affect prognosis. Conversely, although the divergence in likelihood of irAE induction between anti-CTLA-4 and anti-PD1 agents shows that the relationship between anti-tumour effects and irAE development is not linear, our results also suggest that it may be difficult to completely separate efficacy from irAE propensity in novel agents. The comparative strengths of our study are that, unlike data from trials, the results reflect real-world scenarios, with particular relevance to the UK healthcare setting and our observations independently replicate in a separate tertiary centre. Our ability to combine clinical observations with prospectively collected transcriptomic data identified differences in the TCR diversity after treatment according to irAE development, as well as baseline predisposition to activation in sICB recipients who develop irAEs. The limitations of this study include the retrospective collection of clinical data and the relatively small sample size in the primary cohort. Larger transcriptomic series involving other cell subsets will be vital for future studies in teasing out the relationship between clinical response and irAE development. A positive association between increased tumour mutational burden (TMB) and response to ICB is well recognised[30]. Here we show that irAE similarly are positively associated with outcome in MM. Although this likely represents a separate association to that of response and TMB; given the increased baseline inflammatory response in sICB irAE sufferers, it is tempting to speculate that TMB may relate to irAE, with higher TMB eliciting more neo-antigens and potentially off target effects – a hypothesis that warrants further exploration.

In conclusion, in our clinical practice, the development of early irAEs in MM patients treated with ICBs is associated with a survival benefit and, in sICB recipients, development of irAE is associated with baseline immune state. Whether specific irAEs have more favourable prognostic associations than others remains unknown and will require larger datasets to ascertain. These findings highlight the crucial importance of investigating the biological mechanisms underlying irAE development, to determine whether their presence could be a robust indicator of response to treatment. Collectively, this could aid early identification of individuals likely to benefit from ICB therapy, and enable the application of stratified medicine.

## Data Availability Statement

All sequencing data will be made freely available to organizations and researchers to conduct research in accordance with the UK Policy Framework for Health and Social Care Research via a data access agreement. Sequence data will be deposited at the European Genome–phenome Archive, which is hosted by the European Bioinformatics Institute and the Centre for Genomic Regulation under accession no. <u>EGAS00001004081</u>. Patient anonymized irAE data will be shared on reasonable request.

## Data Availability

All sequencing data will be made freely available to organizations and researchers to conduct research in accordance with the UK Policy Framework for Health and Social Care Research via a data access agreement. Sequence data will be deposited at the European Genome Phenome Archive, which is hosted by the European Bioinformatics Institute and the Centre for Genomic Regulation under accession no. EGAS00001004081. Patient anonymized irAE data will be shared on reasonable request.

https://ega-archive.org/search-results.php?query=EGAS00001004081

## Acknowledgements

With thanks to all patients who have generously contributed samples and clinical data to the Oxford Radcliffe Biobank for this study. We are also very grateful to the staff of the Day Treatment Unit at the Oxford Cancer Centre and Brodey Cancer Centre at The Horton Hospital for their invaluable assistance.

## Funding

The work was supported by the National Institute for Health Research (NIHR) Oxford Biomedical Research Centre (BRC). The views expressed are those of the author(s) and not necessarily those of the NHS, the NIHR or the Department of Health. WY is an Academic Foundation Programme Trainee, RAW is an NIHR Academic Clinical Fellow and recipient of a CRUK predoctoral award (C64881/A26189); BPF is funded by a Wellcome Intermediate Clinical Fellowship (201488/Z/16/Z). ACOB is a MRC Academic Clinical Fellow (Award Ref. MR/N025989/1). VTFC is funded by grants from the Norman Collisson Foundation and Oxfordshire Health Services Research Committee (OHSRC) part of Oxford Hospitals Charity

## Disclosures

All authors have declared no conflicts of interest.

## Figures and Tables

**Supplementary Figure 1.**
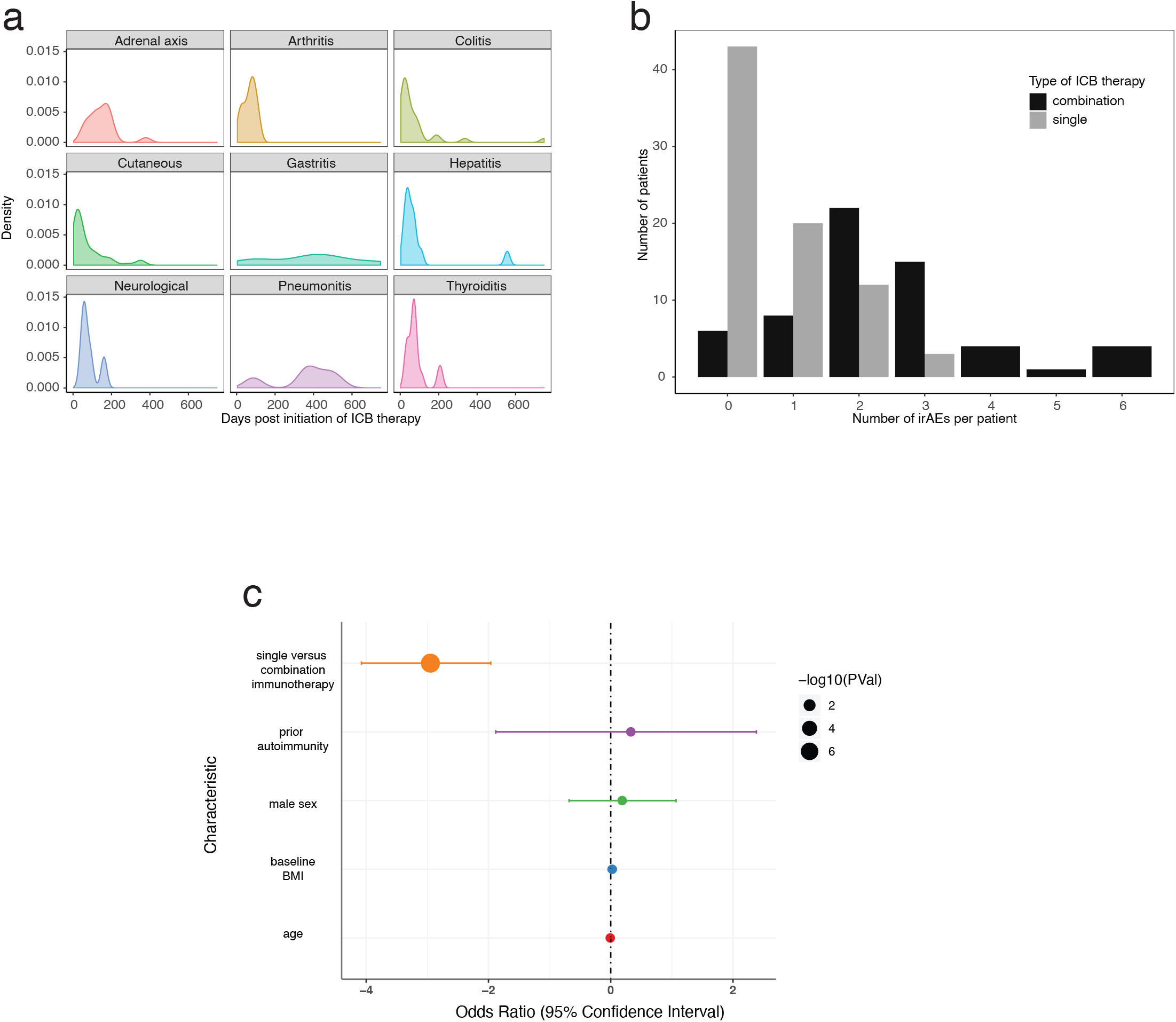
Profile of immune-related adverse events (irAEs) within the cohort. a) Density plot of the development of different irAEs over time. b) Distribution of the number of different irAEs per patient. c) Logistic regression model of the risk factors for development of irAEs prior to cycle 5

**Supplementary Figure 2.**
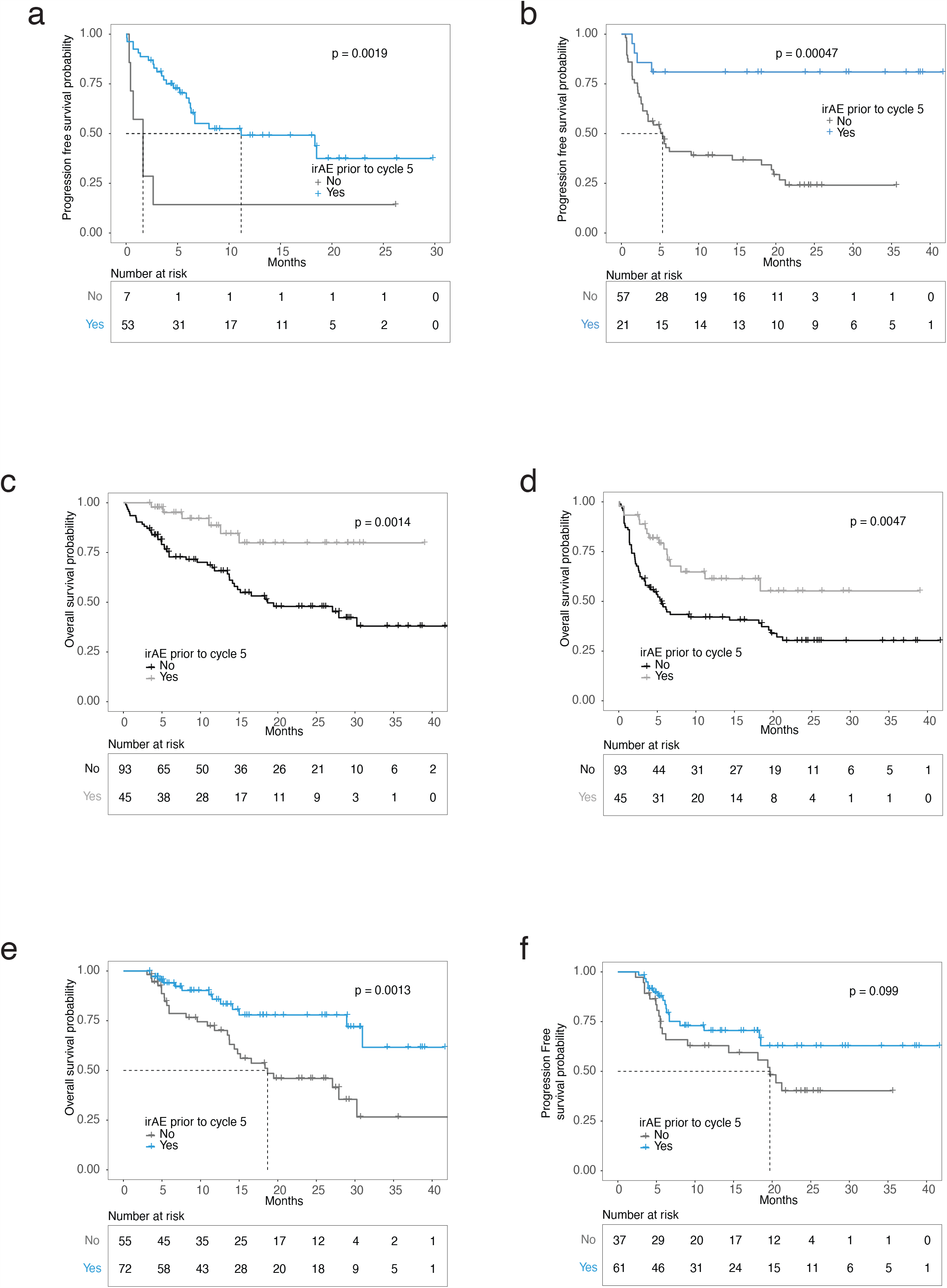
a) Kaplan Meier of Oxford PFS for cICB recipients, b) Kaplan Meier of Oxford PFS for sICB recipients, c) Kaplan Meier of Oxford OS for cICB recipients based on grade 3 or grade 4 toxicity prior to cycle 5, d) as per c, but for PFS, e) Landmark analysis for Oxford OS according to irAE prior to cycle 5, f) Landmark analysis for Oxford PFS according to irAE prior to cycle 5.

**Supplementary Data 1**. List of genes differentially associated with irAE development in pretreatment samples from sICB recipients

**Supplementary Data 2**. List of GOBP pathways enriched for members of genes associated with irAE development

## Notes

### Competing Interest Statement

The authors have declared no competing interest.

### Clinical Protocols

https://ega-archive.org/search-results.php?query=EGAS00001004081

### Author Declarations

Oxford data: Oxford Radcliffe Biobank (09/H0606/5+5), OCHRe 16/A019, 18/A064 Liverpool data: HYST study (12/NW/0525) and local audit approval (17-18/40)

